# Pre-Procedural Surveillance Testing for SARS-CoV-2 in an Asymptomatic Population Shows Low Rates of Positivity

**DOI:** 10.1101/2020.05.08.20078592

**Authors:** James A Mays, Alexander L Greninger, Keith R Jerome, John B Lynch, Patrick C Mathias

## Abstract

Background: Depression is a common cause of mortality and morbidity worldwide. To detect depression, we compared Beck Depression Inventory scoring as a valid tool with participants self-reporting depression.Methodology: This cross-sectional study aimed to explore the diagnostic values of self-reporting in patients' with depression comparing to Beck Depression Inventory scoring in Mazandaran Persian cohort study, with a total of 1300 samples. The sample size was determined to include 155 participants through the census method. In order to increase the test power, 310 healthy participants were included in the study through random selection. In order to evaluate the diagnostic value of self-reporting, BDI-II was completed by blind interviewing to the case group as well as to another group who reported that they were not depressed, as control.

Results: sensitivity, specificity, accuracy, false positive, false negative, positive and negative predictive values of self-reporting was calculated 58.4%, 79.1%,73.4%, 20.8%, 41.6%, 51.8%, and 83.2% for the total population respectively, as well as, sensitivity, specificity, accuracy, positive and negative predictive values of self-report in males were 83.3%, 77.2%, 77.1%, 43.8% and 95.6% and 53.7%, 78.1%, 71.2%, 49.2%, and 81.1% for females, respectively.

Conclusion: The positive predictive value and sensitivity of self-reporting are insufficient in total population and females, and therefore self-reporting cannot detect depressed patients, but regarding to its average positive predictive value, perhaps, it can be used to identify non-depressant individuals.

## Introduction

Seattle region hospitals have been severely impacted for several months by ongoing community spread of the novel coronavirus 2019 disease. (1,2) Although testing was initially focused on the diagnosis and treatment of symptomatic patients, this effort has now expanded to include surveillance of asymptomatic patients in order to protect health care workers and prevent nosocomial infections. There is an urgent need to understand best practices for the delivery of routine medical care during an ongoing outbreak. (3) Here we report the rates of SARS-CoV-2 infection in asymptomatic patients screened prior to admission or a surgical or aerosolizing procedure.

## Methods

Beginning March 30, 2020, our hospital began screening all asymptomatic patients prior to needed surgeries and aerosolizing procedures (n=350). On April 13, 2020, we expanded this practice to universal surveillance screening of all patients prior to admission (n=349). Testing was performed on nasopharyngeal swabs using the Washington state emergency use authorized University of Washington CDC-based laboratory-developed test or FDA authorized DiaSorin Simplexa SARS-CoV-2, Hologic Panther Fusion SARS-CoV-2, or Roche cobas SARS-CoV-2 tests. This study was approved by the Institutional Review Board of University of Washington Medical Center (STUDY00009734). Informed consent was not required.

## Results

For patients undergoing surgical or aerosolizing procedures, 3 of 350 patients (0.9%) were positive for SARS-CoV-2. For patients who were asymptomatic and tested at the time of admission, 3 of 349 patients (0.9%) were positive and 2 of 349 (0.6%) were inconclusive; inconclusive results were treated as low-level positives. For asymptomatic patients tested for any other reason (e.g. exposure risk), 12 of 157 patients (7.6%) were positive and 1 of 157 (0.6%) were inconclusive. Meanwhile, of the 473 inpatients in this period who had any symptom concerning for COVID-19, 68 of 473 patients (14.3%) were positive. There was a variety of ordering services and locations in all indication categories, with no particular predominating medical service or location. During this time period the outpatient prevalence of SARS-CoV-2 active infection in our region was 3–5% (manuscript in review).

## Discussion

The application of universal surveillance testing for SARS-CoV-2 in patients prior to surgery or aerosolizing procedure shows that the positivity rate for SARS-CoV-2 is low (<1%) in asymptomatic patients without known exposure risk factors. The positivity rate for asymptomatic patients on admission screen was similar; both measures were notably lower than recently reported measurements during an outbreak in New York City that found a positivity rate of 13.7% in asymptomatic pregnant women. (4) The rate of sub-clinical infection likely varies with the scale of the community outbreak.

Even in the context of a community-wide outbreak, our data show a low prevalence of COVID-19 infection in the urgent pre-procedural setting. These data reflect the prevalence of asymptomatic SARS-CoV-2 infection in a population of individuals who use medical services and provide an assessment of exposure risk to other patients and healthcare workers around the time of admission. The Greater Seattle Coronavirus Assessment Network (SCAN) also recently published results from the first 18 days of home-based testing and reported no positive tests in 1392 patients reporting no COVID-19-like illness. (5) Although other studies support a large proportion of asymptomatic infections, the data from this metropolitan outbreak do not support a similar pattern. Clinical providers, especially those involved in performing procedures that have a risk of aerosolization, are asking for methods to risk-assess patients prior to the procedure. Pre-procedure testing is one option to accomplish this goal. Importantly, in the midst of an outbreak, testing prior to procedures can decrease the use of PPE, identify appropriate precautions for patients, and reduce the risk of nosocomial infection.

## Data Availability

Data will be made available from the corresponding author by request.

## Acknowledgements

Dr. Greninger discloses personal fees from Abbott Molecular Inc., outside the submitted work.

---

| Indication | Total Patients | Positive (%) | Inconclusive (%) | Negative (%) |
| --- | --- | --- | --- | --- |
| Asymptomatic Admission Surveillance | 349 | 3 (0.9) | 2 (0.6) | 344 (98.6) |
| Asymptomatic (Other) | 157 | 12 (7.6) | 1 (0.6) | 144 (91.7) |
| Pre-procedural Surveillance | 350 | 3 (0.9) | 0 | 347 (99.1) |
| Symptomatic | 473 | 68 (14.3) | 0 | 405 (85.6) |

